# Harmonized Carceral Mortality Database: a dataset on mortality in state-operated correctional facilities

**DOI:** 10.64898/2026.07.29.26359256

**Authors:** Ufuoma Ovienmhada, Gabriela Daza, Robbie M. Parks

## Abstract

Documenting and measuring mortality in correctional facilities is essential for understanding the health consequences of incarceration, identifying preventable deaths, and informing policy interventions. We undertook a compilation of publicly available mortality records in state-operated correctional facilities, encompassing data gathered from administrative, nonprofit, and public records. The Harmonized Carceral Mortality Database (HCMD) standardizes decedent characteristics, facility identifiers, locations, dates of death, and causes of death to enable epidemiologic analyses of mortality in U.S. prisons, outside of federal reporting. At the time of writing, the HCMD includes 49,682 deaths from at least 1,089 prisons in 50 states, spanning 1996 – 2024, with regular updates annually. The HCMD supports carceral health research across a range of disciplines from environmental health to sociology to criminology and supports use beyond the scientific community, including journalists, lawyers, and community-based organizers.

## Background and Summary

Documenting and measuring mortality in correctional facilities is essential for understanding the health consequences of incarceration, identifying preventable deaths, and informing policy interventions. Incarcerated people experience higher rates of chronic medical conditions than the general population ^1^, and each year spent in prison is associated with a two-year decline in life expectancy ^2^. The mortality rate in U.S. state-operated prisons has risen sharply over recent decades: in 2001, the federal government documented a rate of 250 deaths per 100,000 people, compared to350 deaths per 100,000 people in 2018, a 40% increase ^3^. Accurate, systematic data on custodial mortality are therefore critical for public health research, accountability, and the design of interventions to reduce deaths in prisons.

In 2000, Congress passed the Death in Custody Reporting Act (DCRA), which required the Attorney General to collect information on the death of any person who was under arrest, en route to be incarcerated, or incarcerated in a municipal or county jail, state prison, or other local or state correctional facility. To implement this mandate, the Bureau of Justice Statistics (BJS) established the Mortality in Correctional Institutions (MCI) program, which began collecting data from jails in 2000 and state prisons in 2001. BJS achieved an average annual response rate of 98% for local jails and 100% for state prisons ^4^. The MCI collected individual-level data on the number of deaths by year, cause of death, and decedent age, race or Hispanic origin, and sex, enabling detailed comparative analyses across demographic categories, offense types, and facility characteristics.

However, the DCRA of 2000 expired in 2006, and while BJS continued the MCI collection voluntarily, a reauthorized DCRA became law in December 2014. Under the reauthorized law, the Department of Justice determined that the Bureau of Justice Assistance (BJA), rather than BJS, would become responsible for collecting in-custody death data. BJA implemented its data collection plan in October 2019 and BJS published its final MCI report covering deaths through 2019. The transition introduced significant problems: in a review of 2019 data, BJS warned the Department of Justice that BJA’s collection had significant quality and coverage issues, and BJA failed to gather prison death data from 11 states ^5^. BJA has not made any of the data it has collected through the DCRA program publicly available.

Due to these gaps in federal reporting, nonprofits and researchers have developed independent open datasets to fill the void. Non-profit organizations including the Texas Justice Initiative and Incarceration Transparency have long tracked data on deaths in specific states using public records requests. The UCLA Law Behind Bars Data Project, originally the COVID Behind Bars Data Project, built a nationwide database tracking death in U.S. prisons, which documented the 61% surge in prison death rates during the COVID-19 pandemic (https://github.com/uclalawbehindbars/deaths_in_custody). The Third City Project, similarly prompted by the COVID-19 pandemic, started to collect mortality data in 2020 (https://3rdcityproject.com). Together, these efforts represent a growing ecosystem of civic and academic infrastructure attempting to compensate for the collapse of federal transparency.

We present the Harmonized Carceral Mortality Database (HCMD), a harmonized database of deaths occurring in U.S. state-operated correctional facilities, compiled from multiple administrative, nonprofit, and public records. The HCMD standardizes decedent characteristics, facility identifiers, locations, dates of death, and causes of death to enable epidemiologic analyses of mortality in U.S. prisons, outside of federal reporting. The HCMD is the largest publicly available dataset of its kind, extending the scope and usability of both the now-deprecated BJS dataset, which ended in 2019, and the UCLA dataset, which is not georeferenced. The HCMD includes 49,682 deaths from at least 1,089 prisons in 50 states, spanning 1996 – 2024. The database supports carceral health research across a range of disciplines from environmental health to sociology to criminology. The standardization of data across sources enables scaled analyses and makes the database readily accessible to a broader scientific and non-scientific community, including journalists, lawyers, and community-based organizers. Crucially, the HCMD is modular, with supervised and scheduled regular updates to add more data that becomes available or online each calendar year.

## Methods

### Data sources

Information on deaths in correctional facilities were collected from multiple administrative, nonprofit, and public records (**Table 1**). The dataset with the most completeness, the BJS MCI dataset, publicly released records for all 50 states from October 2015 to December 2019 in response to a Freedom of Information Act (FOIA) records request. We browsed through every state department of corrections (DOC) and related government websites for any links to public state-reported individual-level records of mortalities in correctional facilities. We found mortality records directly on the DOC websites for three states (Florida, Montana, Nevada). We found mortality records from non-DOC state agencies for three states (California, Illinois, New York). For each state, we also searched Google for records collated by nonprofit agencies or other non-governmental initiatives using public records requests. We found mortality records from non-governmental sources for four states (Alabama, Louisiana, South Carolina, Texas).

Lastly, we used data from the UCLA Law Behind Bars Data Project () which has collated data of varying temporal coverage for all 50 states using public records requests.

**Table 1.** Sources and coverage of mortality datasets in state-operated correctional facilities.

| Source | Dataset Name | State Coverage | Temporal Coverage |
| --- | --- | --- | --- |
| Bureau of Justice Statistics | Mortality in Correctional Institutions State Prison Data | All 50 states | 10/1/2015 – 12/31/2019 |
| UCLA | Deaths in Custody | All 50 states | 2000 – 2024*, varies by state<br>*Accessed in 2025; new data has since been made available |
| California Department of Justice | Death in Custody & Arrest-Related Deaths | California | 2005 – Present |
| Texas Justice Initiative | Deaths in Custody | Texas | 2005 – Present |
| Alabama Appleseed Center for Law and Justice | Alabama Deaths in Prisons | Alabama | 2022 – 2023 |
| Incarceration Transparency | Louisiana Deaths Behind Bars | Louisiana | 2015 – 2021 |
| Incarceration Transparency | South Carolina Deaths Behind Bars | South Carolina | 2008 – 2024 |
| Illinois Deaths in Custody Project | The Dead 2010 - 2021 | Illinois | 2010 – 2021 |
| Illinois Criminal Justice Information Authority | Death in Custody Reports | Illinois | 2019 – Present |
| Nevada Department of Corrections | Offender in Custody Deaths | Nevada | 2007 – Present |
| Correctional Association of New York | Deaths in Custody | New York | 2020 – Present |
| Montana Department of Corrections | Deaths in Custody | Montana | 2024*<br>*Archived since this project began; the website maintains only same-year published data |
| Florida Department of Corrections | Inmate Mortality | Florida | 2020 – Present |

### Data Compilation

We downloaded datasets from the 13 sources in **Table 1**. When multiple sources provided records for the same state with overlapping temporal coverage, we prioritized state or federal government agency sources over nonprofit or other third-party compilations. When multiple third-party sources were overlapping, we prioritized the dataset with more robust details (e.g. cause of death, demographic statistics). In these cases, we also compared discrepancies and found small differences (e.g. zero death difference for two states, three death difference for one state). We assumed that all discrepancies were valid and manually added any “missing” deaths into the prioritized dataset. To enforce consistent naming conventions, we manually renamed attributes such as decedent name, state, age, sex, and death date across all sources. We split death date information into three columns for death year, death month, and death day. To support further processing at a later step, we also separated attributes into distinct types of prison naming conventions; columns that reported full prison names (e.g. “Cross City Correctional Institution”) were renamed ‘facility name’, whereas columns that reported abbreviated prison names (e.g. “CCCI”) were renamed ‘facility abbrev’. The varied sources reported cause of death information at different levels of specificity (or not at all) ranging from “Illness” to a full paragraph of free text; we differentiated these columns as ‘cause_general’ and ‘cause_specific’.

### Data Standardization

Within Python 3.12.8, our software pipeline integrated all sources into a unified dataset (n = 50,386) and applied several standardization steps to the attribute values. The primary standardization effort targeted ‘facility_name’ and ‘facility_abbrev,’ reported differentially by state. Because most state records omitted direct addresses, these standardized identifier columns served to derive the location of death. We developed a crosswalk of state-specific dictionaries (‘all_facility_dictionaries.py’) to standardize facility names and abbreviations to those used in the Homeland Infrastructure Foundation Level Data (HIFLD) Prison Boundaries shapefile, which is the most comprehensive dataset available of the locations and boundaries of 6,471 U.S. correctional facilities (https://www.arcgis.com/home/item.html?id=70dfcdeedf5243c29738d8c338d55e3f#overview). For example, a single prison might appear as “Cross City C.I.”, “Cross City” or “CCCI” across or even within sources, and the dictionary maps all of those onto the canonical HIFLD name, “Cross City Correctional Institution”.

In Texas, the mortality records did not have facility name or abbreviation information, but they did have a “facility_address” attribute. While these could be used directly as spatial identifiers, some of the addresses reported were postal mailing addresses, not the physical address of the facility where the death occurred. Since a desired use case of this database was environmental health research, we needed to link each decedent to a physical location of death that could be used to infer a collocated environmental exposure at a given facility. Other facility_address values did represent a physical address but used abbreviations or had typos that precluded direct linkage to HIFLD features. Thus, we added address crosswalks to ‘all_facility_dictionaries.py’, linking unique address naming conventions to HIFLD addresses conventions (e.g. “P.O. BOX 9000” linked to “777 FM 3497” for Gib Lewis Prison in Woodville, TX, and “1313 CR 19” linked to “1313 COUNTY RD 19” for Preston E. Smith prison in Lamesa, TX).

We created dictionaries for 49 states that covered a total of 1645 unique naming/address keys linked to the HIFLD name or address. Our goal was not to document every single unique naming convention (nor all unique typos) but to capture a range of colloquial names to enhance the compatibility of the harmonization code with future mortality data from similar and new sources. In states with small DOCs, we manually created the dictionaries. For larger DOCs, we leveraged GitHub Copilot to generate initial key:value predictions (based on patterns in naming conventions by state), then we manually verified each one. State DOC website documents served as the primary source for verifying key-value decodings, while public news media containing the relevant naming conventions provided secondary validation. The decoding references are included as comments in the dictionary to enable scrutiny and transparency.

We implemented a scaffolded, multi-stage join to link a records’ ‘facility name’ or ‘facility abbrev’ to their corresponding HIFLD facility, applying successive matching strategies in order of reliability so that each record was resolved by the most precise method available before falling back to less precise approaches (**Fig. 1**). To allow downstream users to trace and assess linkage confidence for individual records, we tracked the join approach using the variable ‘match_method’. Records that originally came from the UCLA Law Behind Bars Data Project had a unique facility ID that linked to HIFLD based on crosswalks developed and validated in their own project (match_method = ‘cmp_id’, n = 6754). The remaining unmatched records then entered a three-stage, state-by-state facility-name matching process: first, an exact uppercase match between the consolidated facility name and the HIFLD ‘name’ field, restricted to HIFLD records with type == ‘state’ for state-operated prisons (match_method = ‘exact_name’, n = 11285); second, a lookup against the state-specific facility-name crosswalk dictionaries described above, checked against the each original source facility-name or facility-abbrev column in a defined priority order (match_method = ‘name_dict’, n = 22721); and third, fuzzy matching (using a similarity cutoff of 0.76) of the same name fields against the dictionary keys for records that still lacked a match, meant to capture small typos (match_method = ‘fuzzy_name_dict’, n = 1134).

**Fig. 1.**
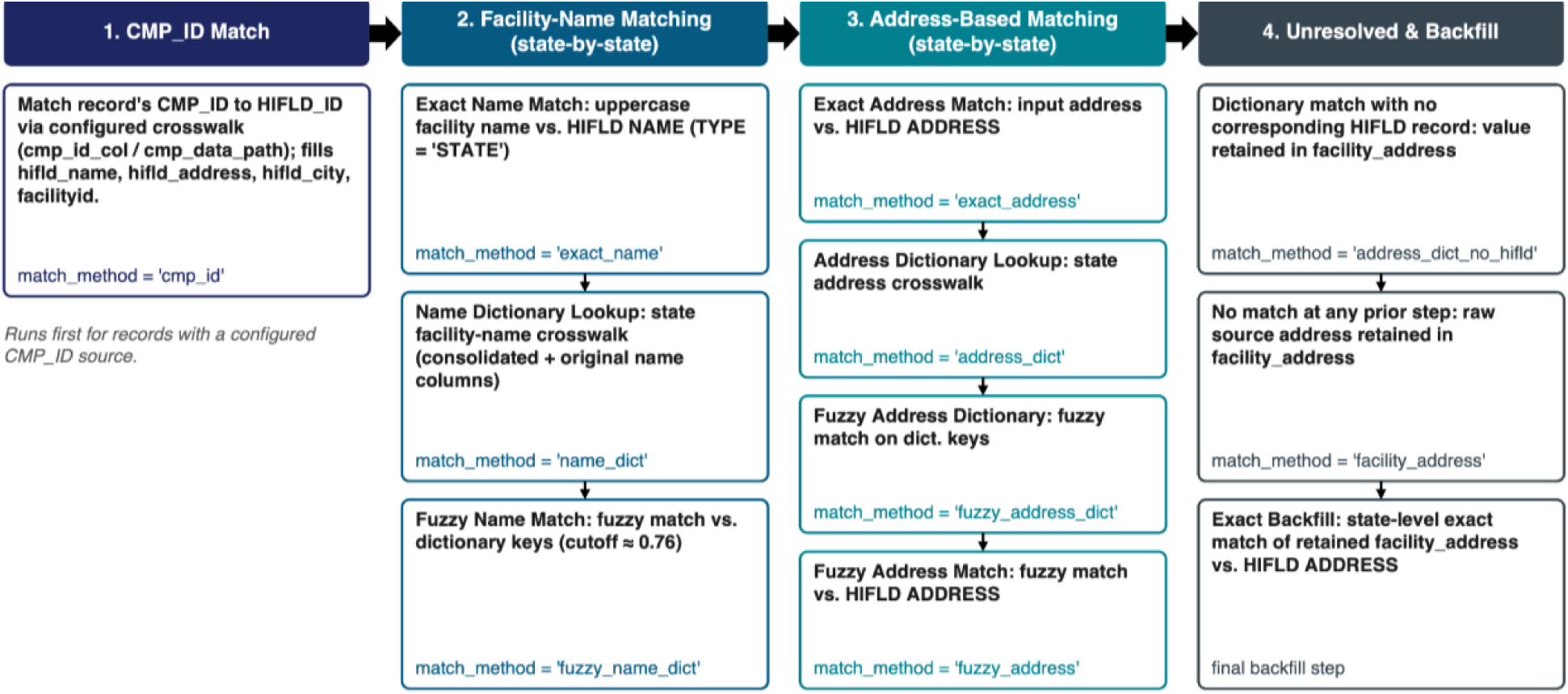
Scaffolded pipeline to link facility identifiers in mortality data to a feature from the Homeland Infrastructure Foundation Level Data (HIFLD) Prison Boundaries dataset. Records pass through successive matching stages, in decreasing order of reliability, until linked to a HIFLD facility record. ‘match_method’ records how each record was resolved.

Records that remained unmatched after name-based linking were then resolved using address information, again through a staged cascade: exact matching of the source address against the HIFLD ‘address’ field (match_method = ‘exact_address’, n = 3205); lookup against state-specific address crosswalk dictionaries (match_method = ‘address_dict’, n = 755); fuzzy matching against the address dictionary keys (match_method = ‘fuzzy_address_dict’, n = 605); and finally, fuzzy matching directly against HIFLD ‘address’ (match_method = ‘fuzzy_address’, n = 190). Records that could not be resolved through any of these steps retained available address information without a confirmed HIFLD linkage: this included cases where an address crosswalk dictionary entry existed but did not correspond to any HIFLD record (match_method = ‘address_dict_no_hifld’, n = 240), as well as records with no dictionary or direct match at all, which retained their raw source address in facility_address (match_method = ‘facility_address’, n = 419). The ‘address_dict_no_hifld’ cases were commonly those where there is a verifiable trace of a facility having existed at an address, but the facility is now closed and no longer in the HIFLD dataset. Lastly, there were some records, exclusively in Texas, where there is no street address reported but there is a city; these records were preserved with match_method = ‘none’ (n = 2374). These latter three match methods precluded linkage to a specific prison, but their geospatial information were still used for spatial information about the location of death for the decedent. Some of the unresolved addresses point to hospitals, presumably in the same city of the prison the decedent originated from; others point to intersections, presumably where a person was being transported in the moments leading up to their death.

Standardization was also applied to unify inconsistent categorical reporting across sources. For example, we developed standardized dictionaries for sex and race/ethnicity, mapping all unique source values onto a coherent, reduced set of categories. Variables for certain decedents were computed on the fly rather than taken directly from source data; for example, age was calculated from the decedent’s date of death and date of birth when both were available. Records were excluded from the final database if they lacked location or spatial data, lacked a recorded date of death, or represented executions rather than deaths from other causes. We excluded 1.3% (n = 665) of records from the original sources and these are preserved in the repository in a file with the prefix “unlinked_…”.

### Data Georeferencing

Following the HIFLD linkage process, records were geocoded to obtain coordinates using a tiered query strategy that used the most precise location information available for each record, informed by the match_method assigned during the HIFLD join. For records with a confirmed HIFLD match, we queried using the HIFLD-derived address, city, and state. For records without a confirmed HIFLD match, we used the best available address information. The variable ‘facility_address’ was used for records with an address that lacked a corresponding HIFLD record. Records with no facility- or address-level information (match_method = ‘none’) were geocoded using city and state alone. In these cases, the geocoded latitude and longitude represent a geocode provider’s best representative point, typically a centroid, of a city.

Geocoding was performed using the Python package geopy, querying two providers in sequence: ArcGIS’s World Geocoding service was queried first; records that could not be resolved by ArcGIS were then submitted sequentially to Nominatim (OpenStreetMap) as a fallback. After applying quality control to the input location identifier data, we successfully geocoded 49,682 records.

### Cause of Death Post-Processing

As noted, we pre-sorted varying levels of cause of death specificity from original records into ‘cause_general’ and ‘cause_specific’ columns. There were originally 1,042 unique ‘cause_general’ and 6,305 unique ‘cause_specific’ string text values. Our goal was to group these columns into a narrower set of ‘cause of death’ values for use in epidemiological analyses compatible and consistent with WHO ICD-10 chapters and causes (https://icd.who.int/browse10/2019/en), while also not providing a false sense of precision given that we were working with relatively sparse information in the original records. We adapted an existing keyword matching algorithm ^6^. The open-source algorithm infers cause of death from free text matched against a lexicon of keywords for eleven cause of death values. We retrieved this algorithm from GitHub and altered it to be best suited for our dataset (**Fig. 2**). In order to maximize the available information for cause of death determination, our amended algorithm preferentially tries to use the “cause_specific” attribute, then the “cause_general” attributes matched against the lexicon of keywords. Further, we amended the lexicon to have keyword sets for four (level 2) broad cause of death values (“Accident”, “Intentional”, “Communicable Disease”, and “Non-communicable disease”) and 14 (level 3) more specific cause of death values (“Cold-related death,” “Heat-related death”, “Overdose”, “Other Accidental”, “Suicide”, “Homicide”, “Other Intentional”, “COVID-19”, “Other Communicable Disease”, “Cancer” “Cardovascular disease” “Respiratory disease”, “Neuropsychiatric conditions”, and “Other Non-Communicable Disease”). Accordingly, we added 69 new keywords to the lexicon that frequently appeared in our dataset, likely due to the carceral specific context. For example, specific terms related to drug use, such as ‘5-flouro-adb toxicity’, were added to the keyword list as indicators of overdose deaths. The output of this algorithm is a csv with ‘true’ or ‘false’ designations in individual columns for each possible cause of death from our lexicon.

**Fig. 2.**
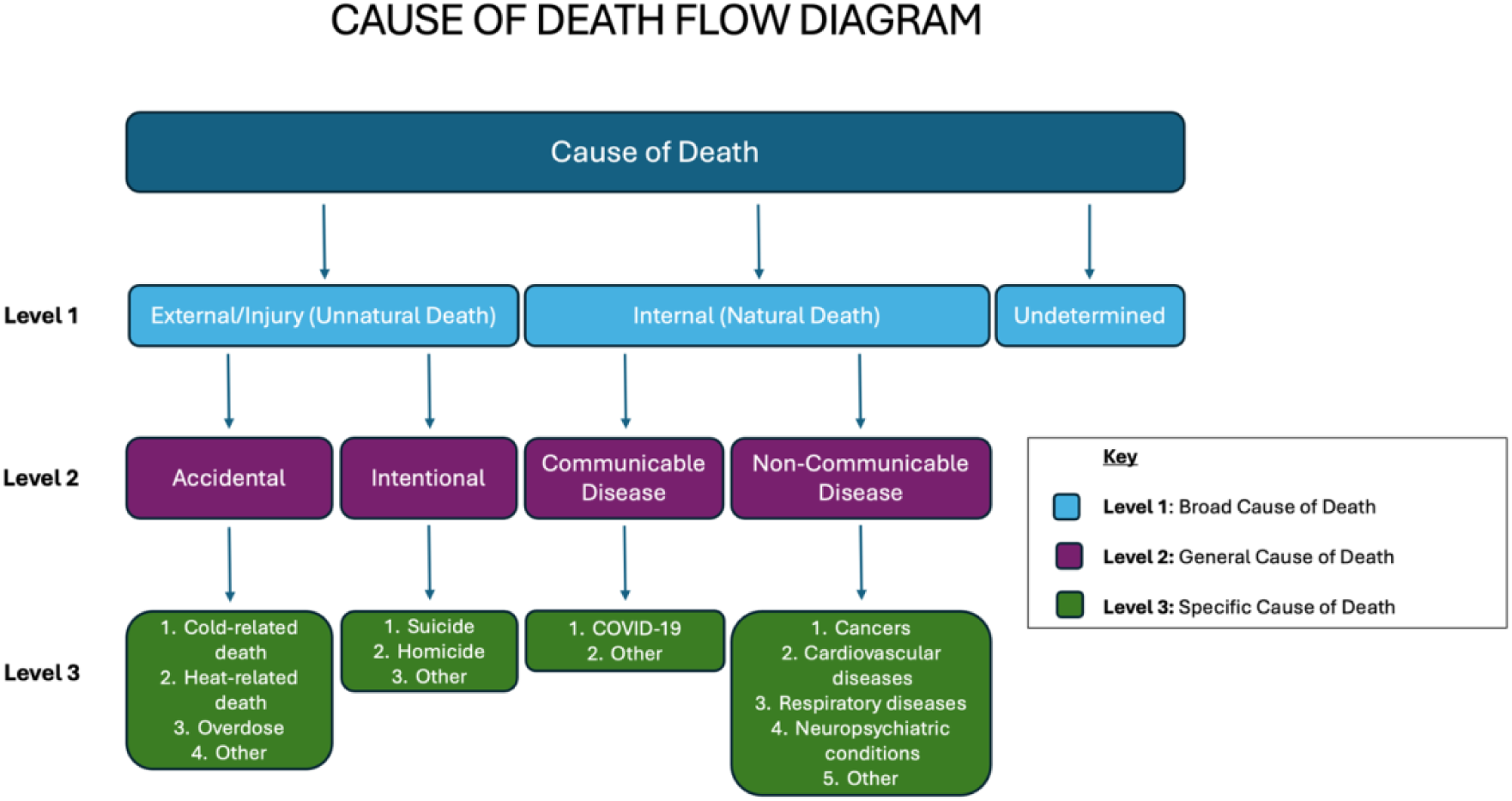
Flow diagram to assign each input decedent to tiered final cause of death categories, with green (level 3) being the highest level of specificity.

We report three ‘level’ attributes that standardize cause of death for each mortality record according to the schema in **Fig. 2**. Our adapted algorithm classifies 42,584 (85.7%) out of 49,682 records with a level_1 classification and 27,189 (54.7%) out of 49,682 with a complete classification across all levels. Examples of two classified records appear in **Table 2**.

**Table 2.**
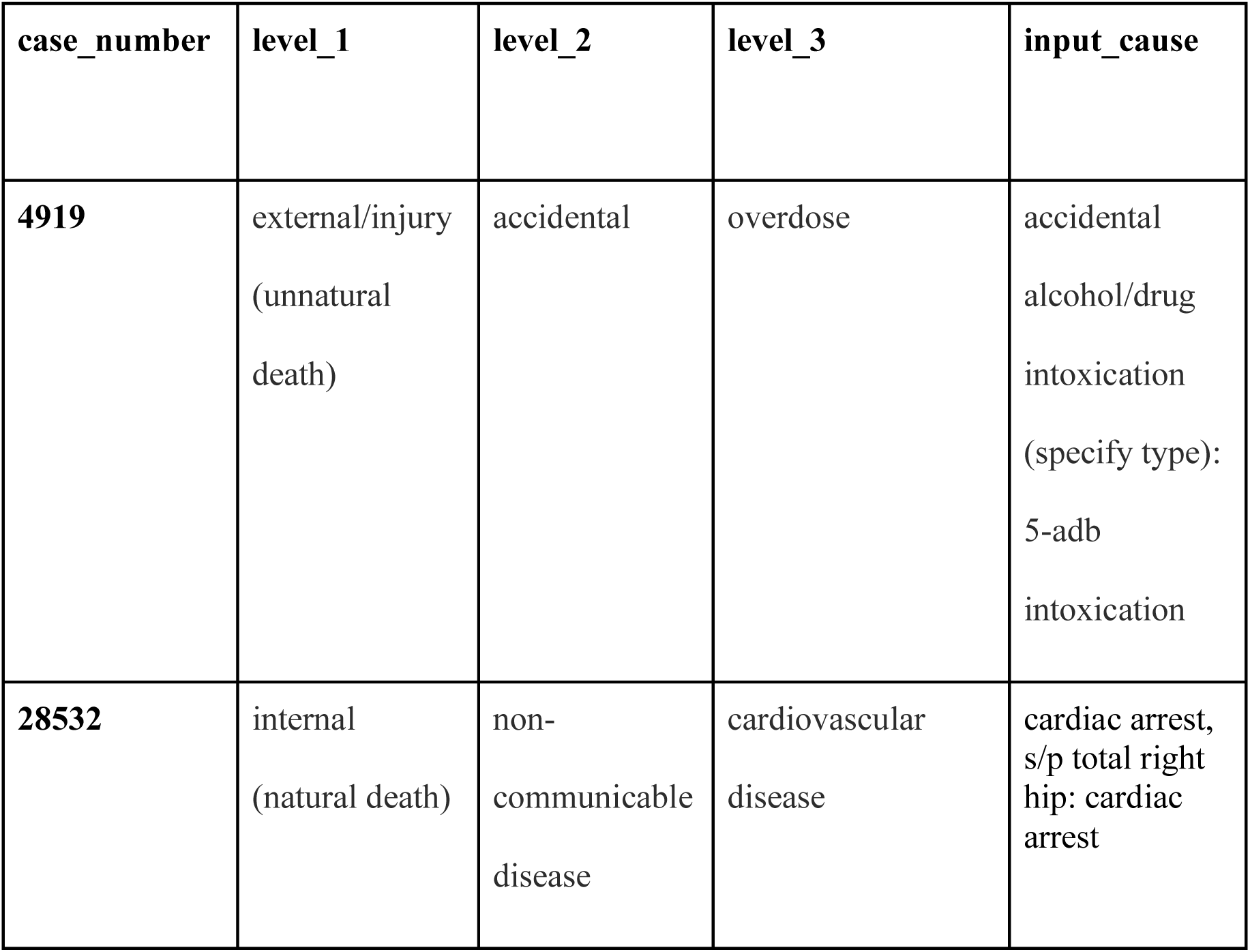
Example input text and classified ‘final cause’ output from algorithm.

## Data Overview

This database contains 49,682 mortality records recorded at varying temporal coverage from March 1996 until December 2024 (**Fig. 3**). Mortality records from all 50 states are included and 1,089 unique carceral facilities are represented in this dataset (**Fig. 4**). Of the 49,682 mortality records, 39,378 records have complete race data. White, non-Hispanic represents 51.0% of the reported population (n = 20,072), followed by Black, non-Hispanic, representing 31.4% of the reported population (n =12,351), and Hispanic or Latino, representing 14.7% of the reported population (n =5,799). Information on sex is available for 80.4% of mortality records (n= 39,944). Females represent 3.7% of the reported population (n=1,479) and males represent 96.3% of the population (n=38,465). Age information is available for 79.9% of mortality records (n= 39,677). People with ages greater than 44 years and ≤ 58 years represent 25.6% of the reported population (n= 12,739), followed by people with ages ≥ 67 years, representing 20.4% of the reported population (n= 10,156), and people ≤ 44 years old representing 20.1% (n= 8,020) of the reported population. People with ages greater than 58 and ≤ 66 years represent the remaining 17.6% of the reported population (n= 8,768). Across the full time period, the climate region ^7^ with the most mortality records was the South (n = 13,060), whereas the Northern Rockies and Plains reported the least amount of mortality records (n = 399). The leading cause of death classification within level 3 across all records that were not undetermined was “*cardiovascular disease”*, representing 25.9% (n= 7,039) (**Fig. 5**).

**Fig. 3.**
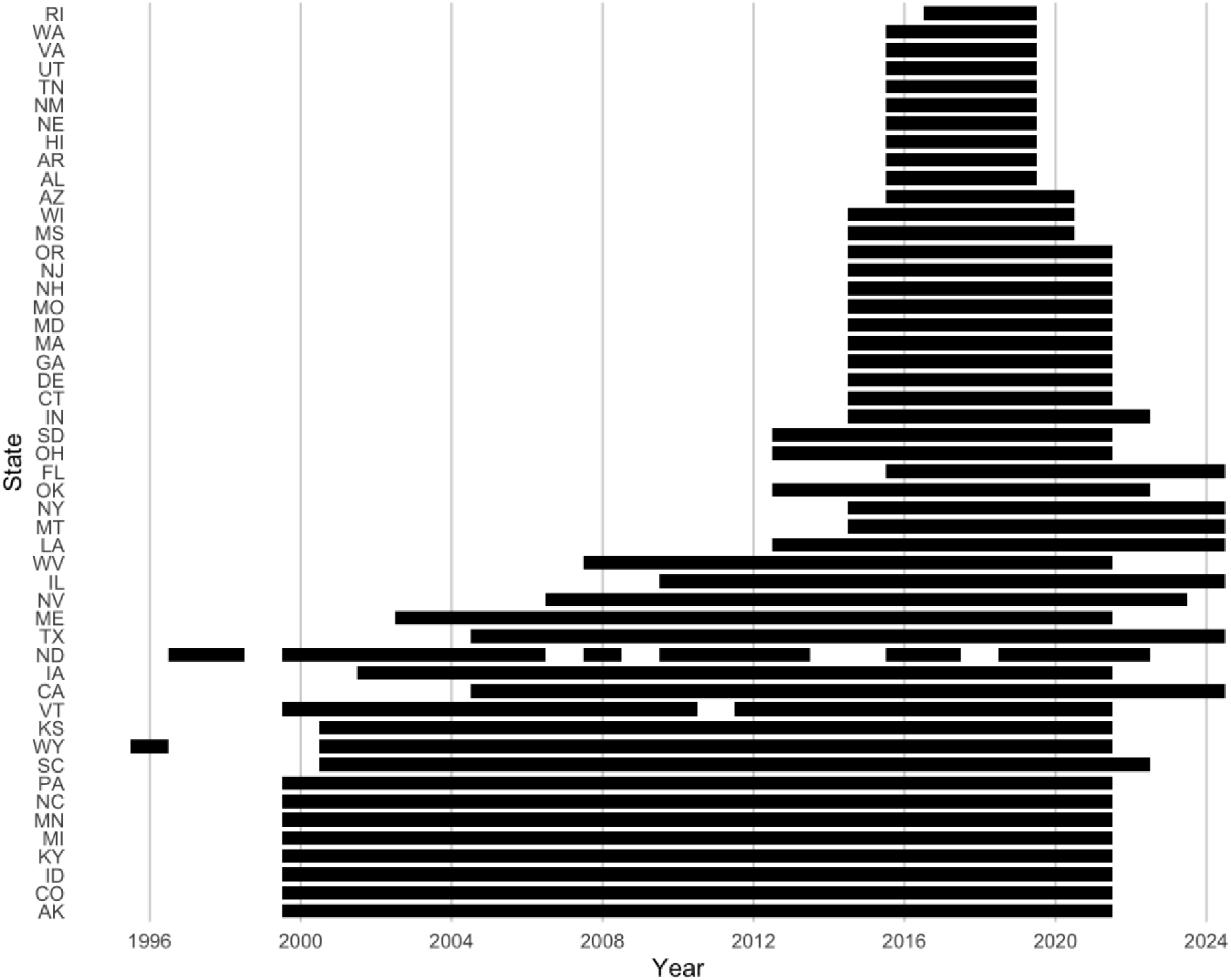
Temporal coverage of incarcerated mortality data by state, 1996-2024, ordered by states with the least to most annual coverage.

**Fig. 4.**
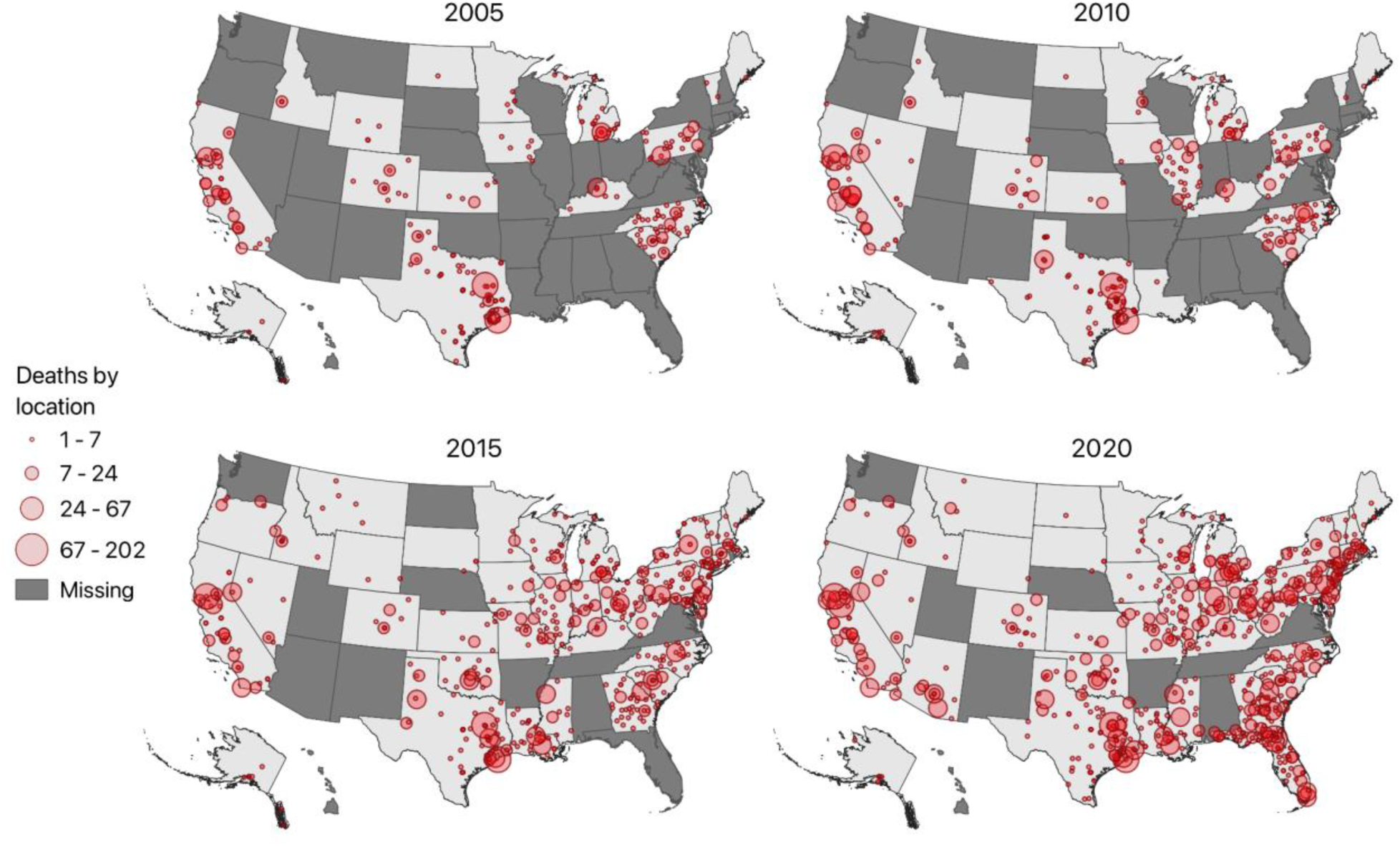
Map of mortality records by location, shown for select years of our full dataset. Each symbol represents a unique location and symbol size corresponds to the number of mortality records at that location in that year.

**Fig. 5.**
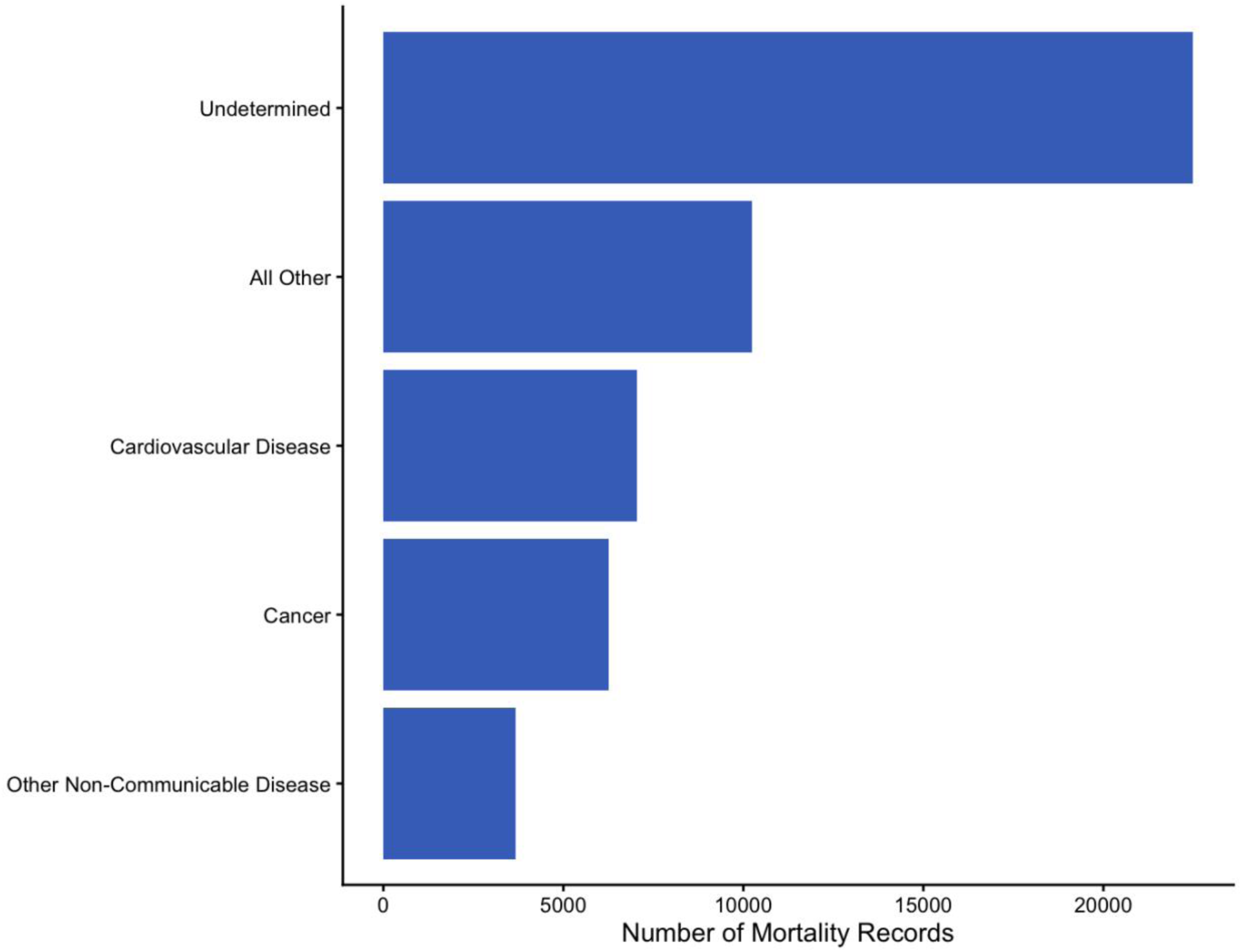
Stacked bar chart showing the top three leading cause of death classifications, all other, and undetermined within the level 3 category assigned by our algorithm.

## Data Records

The HCMD with mortality records in state-operated correctional facilities from 1996 to 2024 and will be available on GitHub under the repository https://github.com/callmeufu/harmonized-carceral-mortality-database. Code files used to harmonize and standardize the data are available on GitHub, and are described in the Code Availability section.

The file, “hcmd.rds”, contains the columns “case_number”, “death_date_dt”, “year”, “death_month”, “state_abbrev”, “state_index”, “latitude”, “longitude”, “match_method”, “hifld_facilityid”, “hifld_name”, “hifld_address”, “hifld_city”, “facility_name”, “facility_address”, “place”, “city”, “location”, “incident_location”, “sex”, “race”, “date_of_birth”, “age”, “age_from_dates”, “class_level_1”, “class_level_2”, “class_level_3”, “matched_keyword”, “cause_general”, “cause_specific”, “illspec”, “drugspec”, “selfspec”, “accspec”, “suicspec”, “homispec”, “otherspec”, “medical_cause”, “days_incarcerated”, “date_incarcerated”.

The column “case_number” is a unique identifier for each decadent. The column “hifld_name” represents the name used by the HIFLD Prison Boundaries dataset to refer to the facility where the death occurred, and “hifld_address” and “hifld_city” refer to the location where each death took place according to HIFLD records. The variables “facility_name”, “facility_address” and “city” refer to the exact facility name, address, and city that appears on the original mortality record. The column “match_method” contains information about how the facility name from each death record was matched to an HIFLD facility. The column “hifld_facilityid” represents a unique identification number, utilized by the HIFLD dataset, for each carceral facility. Variables “latitude” and “longitude” provide geolocations of the place of death. The column “age” provides the age of each individual at their death, if reported on the mortality record. The column “age_from_dates” calculates the age of each individual at death by subtracting “date_of_birth” column information from “death_date_dt” information. The variable “cause_of_death” contains general information about the broad cause or manner of death. The harmonized database combines data from the MCI program as well as state DOC programs. The variables “illspec”, “drugspec”, “selfspec”, “accspec”, “suicspec”, “homispec”, “otherspec”, “place”, “evaluate,” and “cause” contain information about the specific cause of death and are defined by the MCI codebook found on the Department of Justice website (https://www.ojp.gov/program/ojp-freedom-information-act/ojp-reading-room) and our github. The variables “cause_general” and “cause_specific” are standardized columns for any attributes from the original sources in Table 1 that contain information about the cause of death.

## Technical Validation

Review of the harmonization script “a_00_mortality_processing_template.py” was performed by a co-author. Review of the cause of death classification algorithm “a_00_mortality_processing_template.py” was performed by the lead author.

## Usage Notes

This script processes input individual-level carceral mortality datasets and links them to the (now deprecated) HIFLD Prison Boundaries dataset when possible, or other spatial identifiers, to enable more streamlined spatial analyses related to death in custody. It is designed to be generalizable for different data sources while maintaining a consistent output schema. The output dataset has latitudes and longitudes for joining the records to exposure data (e.g. temperature, air pollution). To process new carceral mortality data, users can use existing configuration dictionaries in “a_00_configs.py” or add a new configuration dictionary that identifies the names of the required columns in your specific dataset. The script at minimum requires paths to the unprocessed mortality data, death date column(s), and column(s) indicating facility identifiers (e.g. name, abbreviation). Other columns (e.g. sex, race) are optionally available to configure for standardization according to pre-set dictionary decodings in “a_00_mortality_processing_template.py”. Users then must set “CONFIG_NAME” at the top of “a_00_mortality_processing_template.py” to indicate which configuration should be used. If linkage performance is poor, consider adding new key:value pairs to “all_facility_dictionaries.py”.

Users should be aware of several limitations of the HCMD:

- **Geographic mortality data representation:** There is an evident bias in the distribution of records. Data was available primarily for the South (26.3% of records), Southeast (19.6%), the West (16.4%), Northeast (12.3%), and Ohio Valley (12.3%). Conversely, there is a lower proportion of records from the Southwest (3.5%), Upper Midwest (7.0%), Northwest (1.5%), and Northern Rockies and Plains (0.8%). Some of this distribution is expected (e.g. Texas which is in the Southern region has the highest incarcerated population in the United States) whereas some of the distribution is simply an artifact of which states have publicly available mortality data that we obtained for the HCMD, which may not necessarily correspond to the prisons where most deaths in state-operated correctional facilities actually occur.
- **Location Accuracy:** Unique facility identifiers from different sources were decoded and crosswalked to facilities in the HIFLD Prison Boundaries dataset to the best of our ability but some of these decodings (in particular, of abbreviations) could be wrong. Further, some records listed multiple locations (seen in Kentucky), or hospitals (Wisconsin), presumably where someone may have been transferred before being pronounced deceased. All assumptions in the facility location decoding or logged as comments in “all_facility_dictionaries.py” enabling traceability of decision making and particularly ambiguous cases
- **Cause of Death Accuracy:** 21.7% of records (n= 10,771) lacked any cause of death information and 64.0% (n= 31,785) only included very broad information (e.g. “Natural”). Our algorithmic cause of death classification is thus partial, biased, and should be understood as cautiously descriptive of broad cause of death

Acknowledging these limitations, it is important to highlight that the HCMD is the largest publicly available dataset of its kind, enabling more accessible awareness and analyses of the occurrence and location of deaths in state-operated correctional facilities. Future updates will be made annually in the summer to integrate new mortality data from the previous year. New versions will be published under new unique identifiers.

## Data Availability

The HCMD dataset containing mortality records in state-operated correctional facilities from 1996 to 2024 will be available on GitHub under the repository https://github.com/callmeufu/harmonized-carceral-mortality-database.

## Code Availability

The Python code used to process the HCMD will be available at https://github.com/callmeufu/harmonized-carceral-mortality-database.

## Notes

### Competing Interest Statement

The authors have declared no competing interest.

### Author Declarations

This is a dataset paper documenting the creation of a new database of mortality in correctional settings. Data was obtained exclusively from publicly available sources including administrative, nonprofit, and public records requests (performed by other entities). A full list of source datasets and their coverage is documented in Table 1 of our paper and in the README.md of the private repository associated with this paper. Upon publication in a peer-reviewed journal, a public repository with the table and links to original sources will be made available.

## References

1. Maruschak, L. Medical Problems of State and Federal Prisoners and Jail Inmates, 2011–12. (2016).

2. Patterson, E. J. The Dose–Response of Time Served in Prison on Mortality: New York State, 1989–2003. Am J Public Health 103, 523–528 (2013).

3. Carson, E. A. Mortality in State and Federal Prisons, 2001–2019 – Statistical Tables. (2021).

4. National Academies of Sciences, E., Affairs, P. and G., Committee on Science, T. & Investigations, C. on A. the F. of F. P. L. L. from D.-C. Data on Deaths and Deaths in Custody. in Strengthening the U.S. Medicolegal Death Investigation System: Lessons from Deaths in Custody (National Academies Press (US), 2025).

5. Ossoff, J. & Johnson, R. UNCOUNTED DEATHS IN AMERICA’S PRISONS & JAILS: HOW THE DEPARTMENT OF JUSTICE FAILED TO IMPLEMENT THE DEATH IN CUSTODY REPORTING ACT. https://www.hsgac.senate.gov/wp-content/uploads/imo/media/doc/2022-09-20%20PSI%20Staff%20Report%20-%20Uncounted%20Deaths%20in%20America’s%20Prisons%20and%20Jails.pdf (2022).

6. McCord, K. et al. Algorithm Development for the Automation of Death Certificate Analysis and Coding. Ann Epidemiol 104, 15–20 (2025).

7. Karl, T. Regional and National Monthly, Seasonal, and Annual Temperature Weighted by Area, 1895-1983. (National Climatic Data Center, Asheville, North Carolina, 1984).

